# Enhanced neutrophil extracellular trap formation in COVID-19 is inhibited by the PKC inhibitor ruboxistaurin

**DOI:** 10.1101/2021.08.24.21262336

**Authors:** Rebecca Dowey, Joby Cole, A A Roger Thompson, Chenghao Huang, Jacob Whatmore, Ahmed Iqbal, Kirsty L Bradley, Joanne McKenzie, Rebecca C Hull, Allan Lawrie, Alison M Condliffe, Endre Kiss-Toth, Ian Sabroe, Lynne R Prince

## Abstract

Neutrophil extracellular traps (NETs) are web-like DNA and protein lattices which are expelled by neutrophils to trap and kill pathogens, but which cause significant damage to the host tissue. NETs have emerged as critical mediators of lung damage, inflammation and thrombosis in COVID-19 and other diseases, but there are no therapeutics to prevent or reduce NETs that are available to patients. Here, we show that neutrophils isolated from hospitalised patients with COVID-19 produce significantly more NETs in response to LPS compared to cells from healthy control subjects. A subset of patients were captured at follow-up clinics (3-4 month post-infection) and while LPS-induced NET formation is significantly lower at this time point, it remains elevated compared to healthy controls. LPS- and PMA-induced NETs were significantly inhibited by the protein kinase C (PKC) inhibitor ruboxistaurin. Ruboxistaurin-mediated inhibition of NETs in healthy neutrophils reduces NET-induced epithelial cell death. Our findings suggest ruboxistaurin could reduce proinflammatory and tissue-damaging consequences of neutrophils during disease, and since it has completed phase III trials for other indications without safety concerns, it is a promising and novel therapeutic strategy for COVID-19.

## Introduction

Excessive inflammation is characteristic of severe COVID-19 disease. Neutrophils are recruited to the lungs in response to SARS-CoV-2 infection and are a principal cause of tissue damage and ongoing inflammation (1). Neutrophil activation at the alveolar space is thought to contribute to the development of acute respiratory distress syndrome (ARDS) in COVID-19 as well in other lung infections (2-4). Here, neutrophils perform antimicrobial effector functions including production of reactive oxygen species (ROS), degranulation of cytotoxic proteins and release of NETs via NETosis. NETs are extracellular DNA lattices coated in histones and antimicrobial proteins including cathepsins and myeloperoxidase (MPO). NETs are antimicrobial, but they also cause significant host tissue damage and exacerbate inflammation in multiple acute and chronic diseases, including those of the lung (5). NET production is increased during COVID-19, with NETs identified in both lung autopsy samples from deceased patients with COVID-19 and in bronchoalveolar lavage fluid (BALF) (6-8). Furthermore, SARS-CoV-2 directly induces NETosis *in vitro*, via a ROS dependant mechanism, and circulating markers of NETosis (including cell free DNA and NE) are associated with increased COVID-19 severity (6, 7, 9, 10). NETs are highly pro-thrombotic *in vivo*, aggregating with platelets and the activated endothelium in COVID-19, to form microthrombi which occlude the vasculature and further perpetuate inflammation (6). Furthermore, SARS-CoV-2 induced NETs induce epithelial cell death, driving the catastrophic damage to the airway epithelium that is associated with severe disease (8). This growing evidence indicates that inhibiting NET formation is an important and viable therapeutic strategy. Here we show for the first time that NETs are elevated in response to LPS from neutrophils isolated from hospitalised patients with COVID-19 and that the orally active PKC inhibitor, ruboxistaurin (LY-333531), is a potent inhibitor of NETosis in this cohort. Since ruboxistaurin has completed phase III trials for other indications and is safe in humans, we believe it could be quick to enter the clinic as a new drug for COVID-19.

## Results

### Neutrophils isolated from patients with COVID-19 generate more NETs in response to LPS

Neutrophils were isolated from venous blood from healthy volunteers (healthy controls) or patients hospitalised following a positive PCR test for SARS-CoV-2 (n=39). Of the 39 COVID-19 patients recruited to the study, 38 required supplemental O_2_, 32 received dexamethasone, 3 were subsequently admitted to intensive care, and 2 died (Table 1). Neutrophils were treated with LPS or phorbol myristate acetate (PMA), which induce NADPH oxidase- and PKC-dependent NETosis (11-14). NET formation was measured by SYTOX™ Green staining of extracellular DNA (9, 13). Compared with healthy control subjects, neutrophils from people with acute COVID-19 generated significantly more NETs in response to LPS and a similar amount of NETs in response to PMA (Figure 1). Three patients were admitted to the intensive care unit (ITU) during our study (indicated as open red squares, Figure 1) and generated among the highest SYTOX green values following PMA treatment. The increased capacity of neutrophils to undergo LPS-induced NETosis during the acute stage of COVID-19 adds to existing data suggesting this could be a key element of the dysregulated and deleterious inflammatory response in COVID-19.

**Table 1.** COVID-19 patient characteristics

|  |  |
| --- | --- |
| **Demographics** |  |
| Total number of participants | 39 |
| Age in years: mean ± stdev | 57.4±12.3 |
| Age in years: range | 29-83 |
| Female: number (percent) | 12 (30.8%) |
| Male: number (percent) | 27 (69.2%) |
| **Clinical data** |  |
| Days following symptom onset of neutrophil sampling: mean±stdev | 12.9±7 |
| Days following symptom onset of neutrophil sampling : range | 4-43 |
| Length of stay in hospital (days): mean±stdev | 11±13.2 |
| WHO symptom severity score: mode | 1 |
| Required supplemental O2: number (percent) | 38 (97.4%) |
| Receiving dexamethasone: number (percent) | 32 (82.1%) |
| Receiving tocilizumab: number (percent) | 1 (2.6%) |
| Admitted to ITU: number (percent) | 3 (7.7%) |
| Deaths: number (percent) | 2 (5.1%) |
| **Co-morbidities: number (percent)** |  |
| Diabetes (incl. pre-diabetes) | 14 (35.8%) |
| Hypertension | 11 (28.2%) |
| Asthma | 9 (23.1%) |
| Cancer | 6 (15.4%) |
| Cardiovascular Disease | 5 (12.8%) |
| Obesity | 4 (10.3%) |
| Kidney Disease | 2 (5.1%) |
| COPD | 2 (5.1%) |
| Bronchiectasis | 1 (2.6%) |

**Figure 1.**
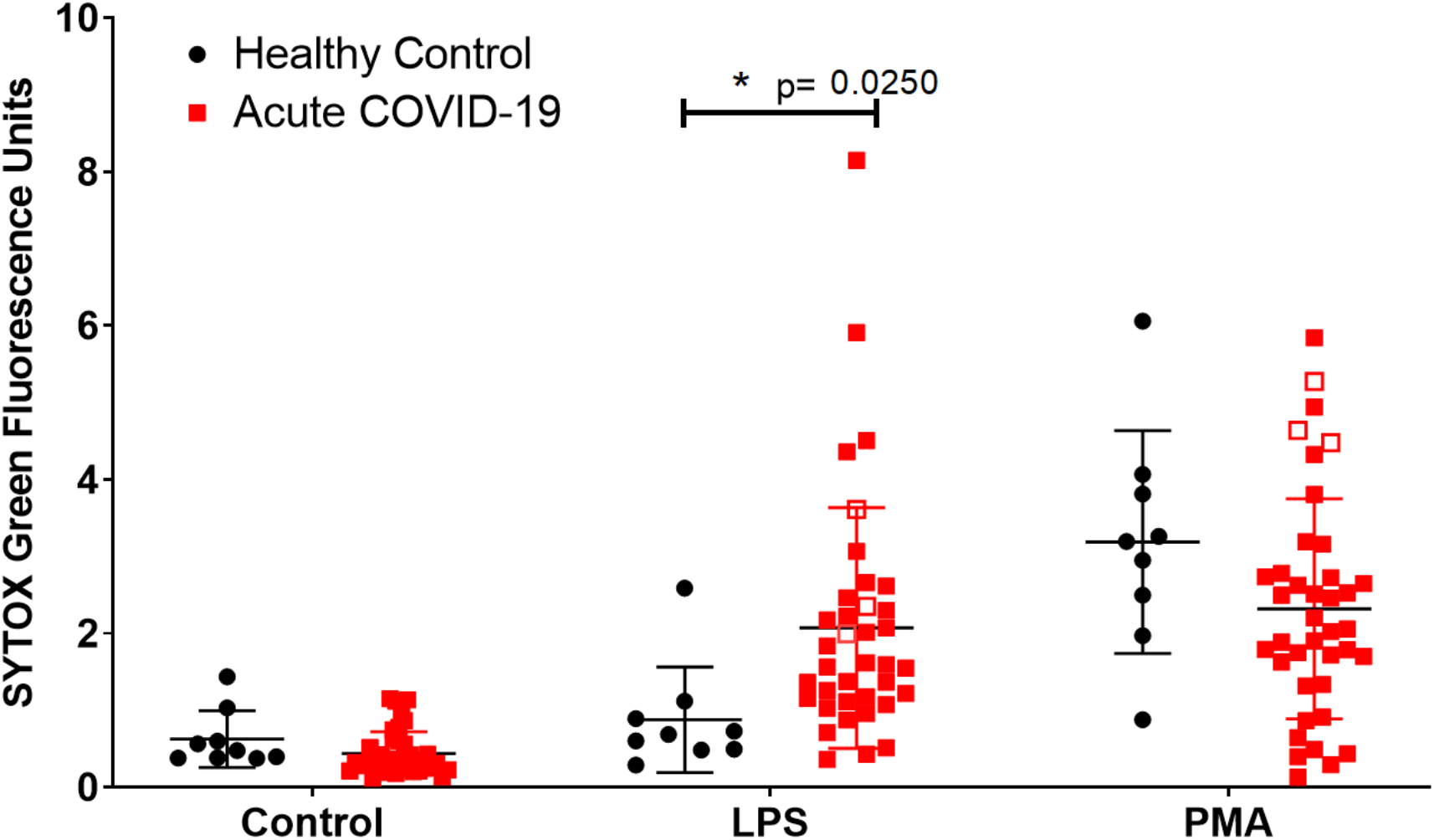
LPS stimulated NET release is significantly elevated in acute COVID-19 patients. Neutrophils isolated from peripheral whole blood from healthy control subjects (black circles, n=9) or hospitalised patients with COVID-19 (red square, n=37 LPS, n=39 PMA), were stimulated for 3 hours with either LPS [5 µg/ml] or PMA [100 nM]. Open red squares indicate patients (n=3) who were admitted to ITU during the study. SYTOX green was added, and extracellular DNA release (NETs) was quantified using a fluorescent plate reader. A significant increase in NET formation was shown in acute COVID-19 patients in response to LPS but not in response to PMA. Statistical analysis used a mixed-effects model with a Šidák post-test. Error bars represent standard deviation.

### The orally active inhibitor of PKC, ruboxistaurin, inhibits LPS-induced *ex vivo* NET formation in COVID-19

NETosis can occur via ROS-dependent mechanisms (10, 15, 16). In support of this, we show both PMA- (Figure 2A) and LPS- (Figure 2B) induced NET formation in neutrophils from people with acute COVID-19 is significantly reduced by the NADPH oxidase inhibitor, diphenyleneiodonium (DPI). PKC is a key signalling component of ROS-dependant NET formation (13). Ruboxistaurin is an effective inhibitor of PKC-β, has completed phase III clinical trials for diabetic retinopathy and is well-tolerated by patients (17). We show for the first time that ruboxistaurin is a potent inhibitor of NET formation in COVID-19 neutrophils, significantly reducing both LPS- (Figure 3A) and PMA- (Figure 3B) induced NETs. During NETosis, neutrophils release DNA which is decorated with antimicrobial components including myeloperoxidase (MPO) (18). We confirm biochemically and morphologically that neutrophils from patients with COVID-19 generate MPO-positive NETs in response to PMA and LPS and that fewer NETs are visualised in the presence of ruboxistaurin (Figure 3C). NETs directly induce epithelial cell damage (8, 19). Here we show that supernatants (SPNs) from PMA-treated neutrophils isolated from healthy volunteers induce death of human bronchial epithelial (HBEC3-KT) cells, which is significantly reduced by ruboxistaurin (Figure 4A-B). Rounding up and detachment of the monolayer was visible in HBEC3-KT cells cultured with SPNs from PMA treated neutrophils, which was reduced in the presence of ruboxistaurin (Figure 4C). Since secondary infections are not uncommon in COVID-19, we also measured killing of the human pathogen *Staphylococcus aureus* by COVID-19 neutrophils and show ruboxistaurin had no effect on the ability of neutrophils to kill S. *aureus* (data not shown).

**Figure 2.**
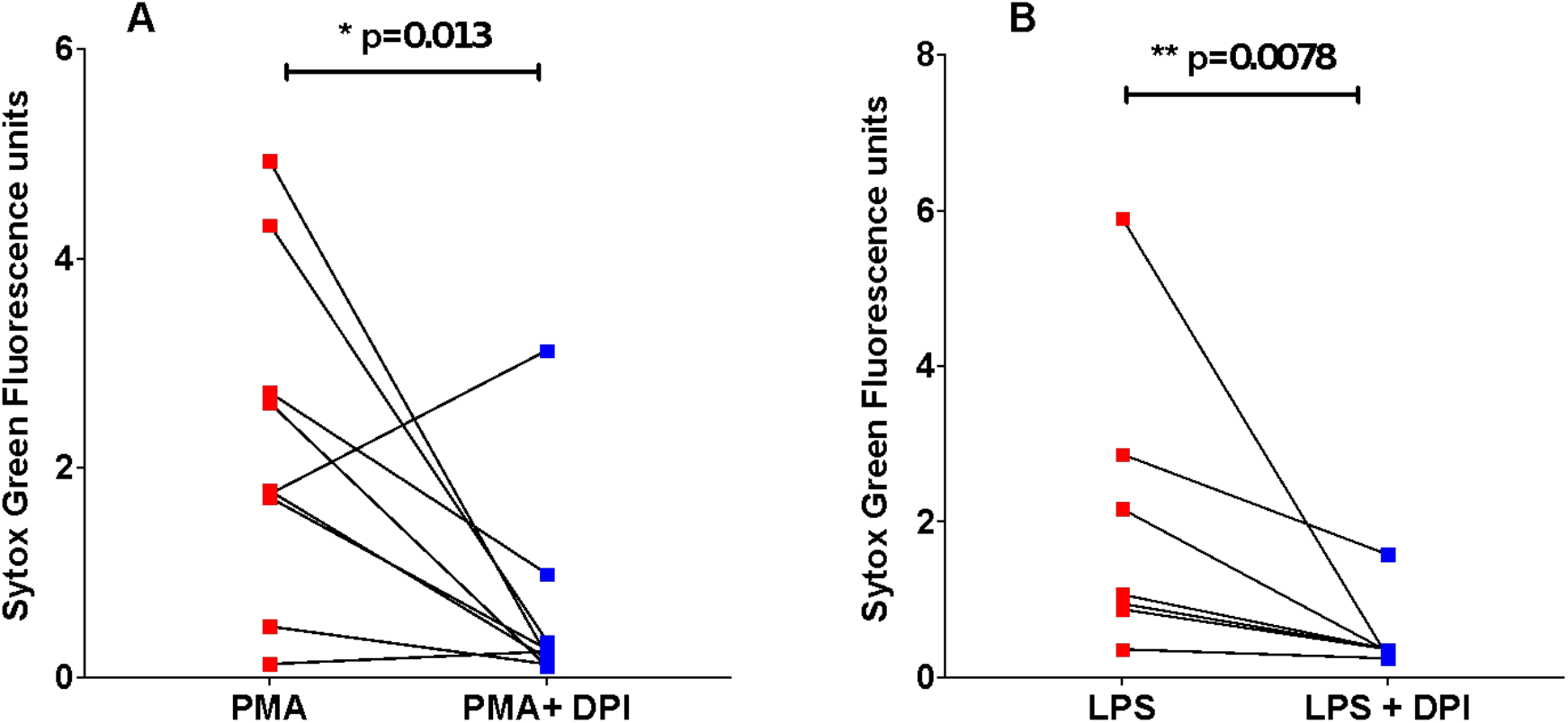
The ROS inhibitor DPI significantly reduces both PMA- and LPS-stimulated NET formation in neutrophils from acute COVID-19 patients. Neutrophils isolated from peripheral whole blood from hospitalised patients with COVID-19 were pre-incubated with ROS inhibitor, DPI (10 µM), for 1 hour (blue squares). Neutrophils were stimulated with PMA [100 nM] (A, n=9) or LPS [5 µg/ml] (B, n=7) for a further 3 hours (red squares). SYTOX Green was added and extracellular DNA (NETs) was quantified using a fluorescent plate reader. Statistical analysis was performed by one-tailed Wilcoxon matched-pairs signed rank test and significance values are as indicated.

**Figure 3.**
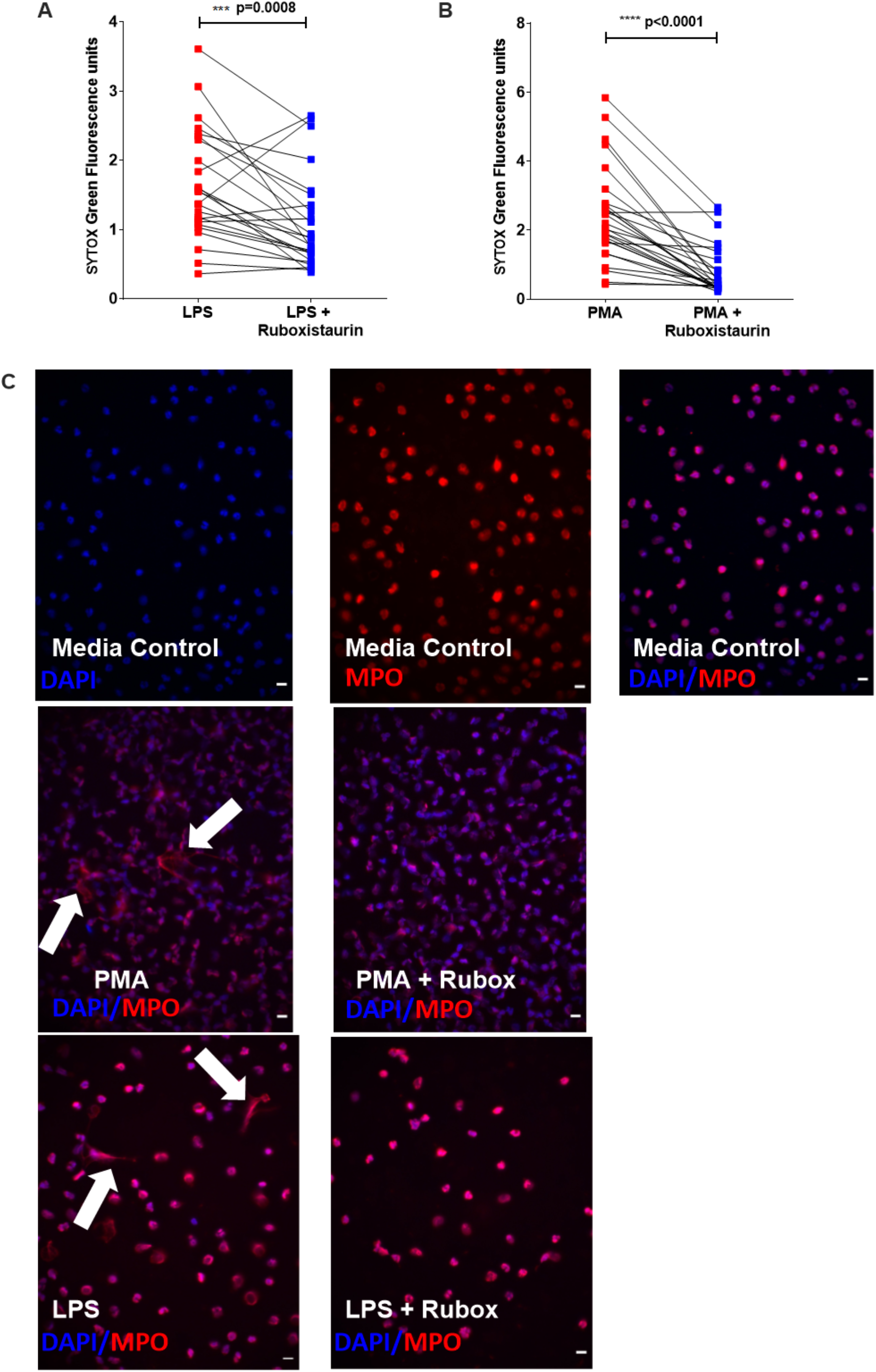
Ruboxistaurin significantly reduces both LPS- and PMA-stimulated NET formation in neutrophils from acute COVID-19 patients. Neutrophils isolated from peripheral whole blood from hospitalised patients with COVID-19 (A n=26, B n=28) were preincubated with ruboxistaurin [200 nM] for 1 hour (blue squares). Neutrophils were stimulated with LPS [5 µg/ml] or PMA [100 nM] for 3 hours (red squares). SYTOX green was added and extracellular DNA (NETs) was quantified using a fluorescent plate reader. Statistical analysis was performed by Wilcoxon matched-pairs signed rank test (A, B) and significance values are as indicated. (C) COVID-19 patient derived neutrophils were seeded in IBIDI(tm) chamber wells and stimulated as described for panels A-B, plus media control. Neutrophils were stained for myeloperoxidase (MPO) and detected using Alexafluor 597 fluorochrome (red). DNA was visualised with ProLong(tm) Gold Antifade Mountant with DAPI (blue). Cells were viewed by fluorescence microscopy (40x magnification) and images are representative of 3 independent experiments. Fields of view were selected at random. Arrows indicate NETs. Scale bar = 10 µm.

**Figure 4.**
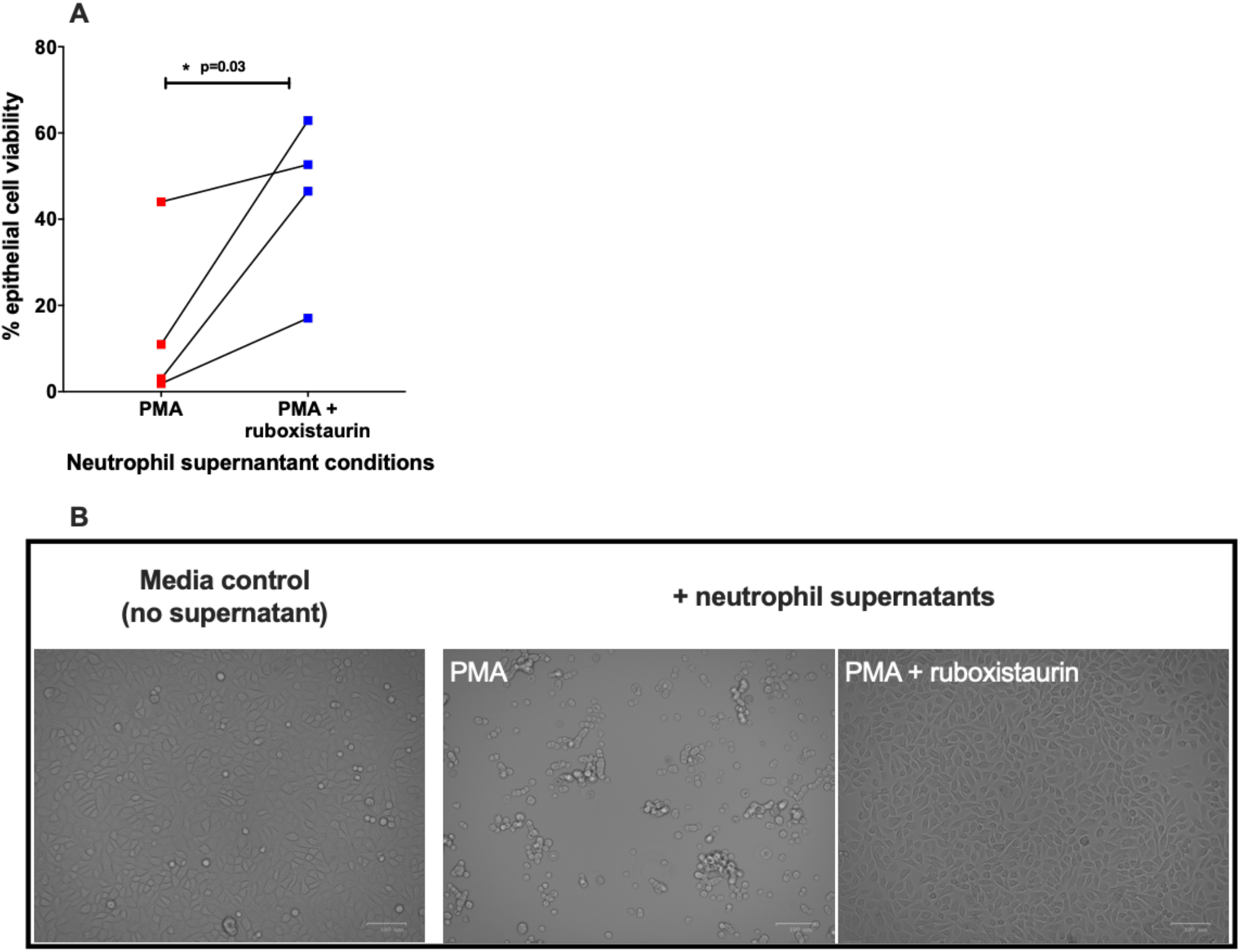
Ruboxistaurin reduces neutrophil supernatant-induced epithelial cell damage. Cell free SPNs from neutrophils isolated from healthy donors stimulated with PMA [100 nM] ± ruboxistaurin [200 nM] were added to confluent human bronchial epithelial cells (HBEC3-KT) at a 1:2 dilution (A) After 24 hours HBEC3-KT cell viability was assessed using CellTitre Glo^®^(n=4). B) Cell monolayers were imaged using the Zoe fluorescent cell imager, using the brightfield setting and 20x objective lens. Images are representative of 4 donors and fields of view were selected at random. Scale bar represents 100 µm. Statistical analysis was performed by Student’s t-test and significance values are as indicated.

### Elevated NETosis in acute COVID-19 patients reduces over time but remains higher than in healthy controls

Neutrophils have to shown to be reprogrammed during COVID-19 and we aimed to investigate whether the pro-NET phenotype observed in our acute cohort persisted post infection (20). To do this we studied a subset of 7 individuals at follow-up clinics held 3 - 4.5 months post hospitalisation. LPS-induced NETosis was significantly reduced at the follow-up time point (Figure 5A) indicating a reduction in the pro-NET phenotype in this population. PMA-induced NETosis did not differ between the acute and follow-up time points (Figure 5B). This is not unexpected since PMA is a potent inducer of NETs in healthy cells. In comparison to NETosis in healthy neutrophils however, (previously shown in Figure 1A) LPS-induced NET formation (Figure 5C) but not PMA-induced NET formation (Figure 5D) remained significantly elevated in neutrophils isolated at the follow-up time point.

**Figure 5.**
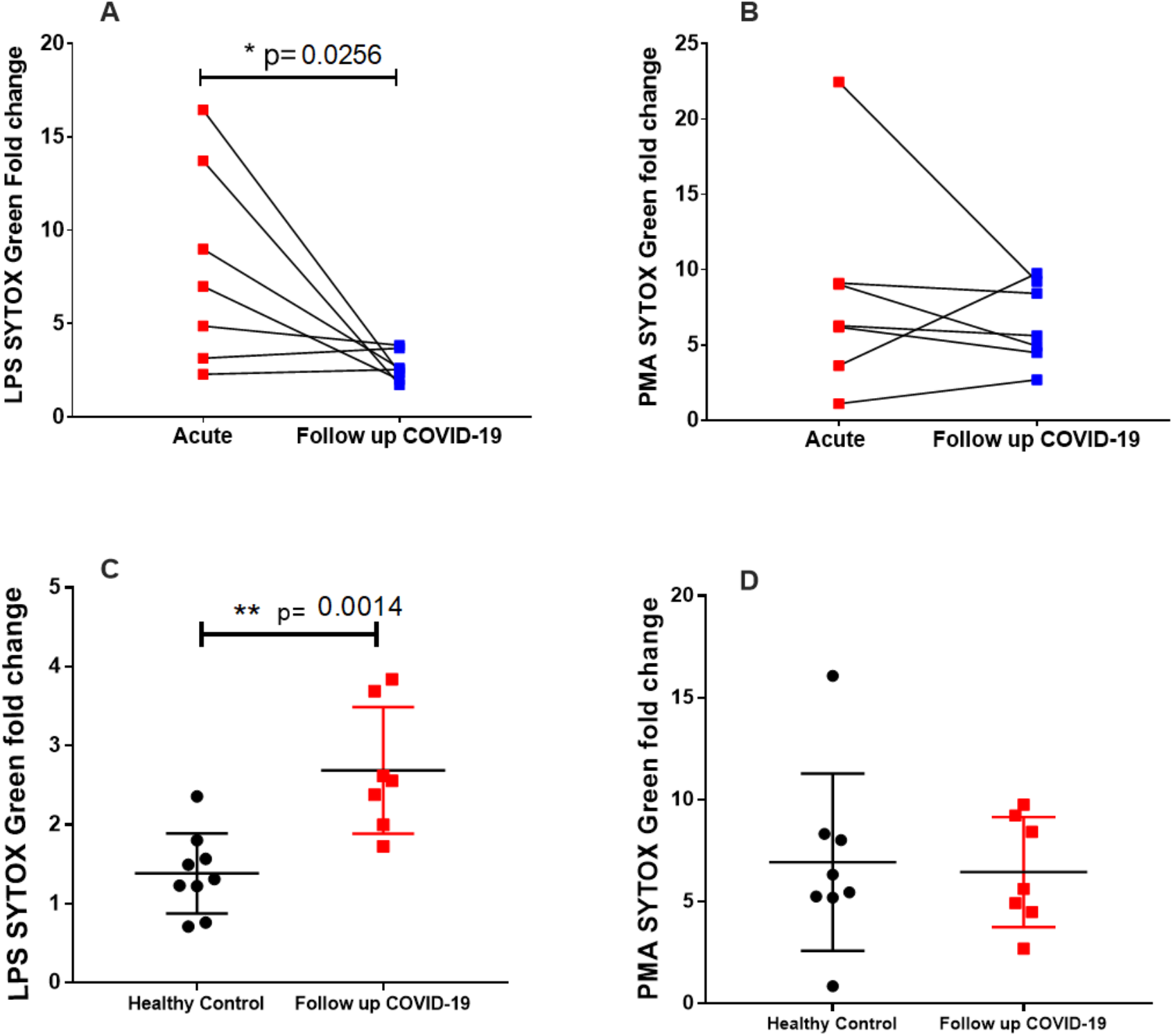
LPS-induced NETosis is reduced in follow-up COVID-19 patients but remains significantly higher than in healthy controls. Seven previously hospitalised patients with COVID-19, who were part of the acute COVID-19 cohort in Figure 1A, returned to a follow up clinic 3-4 months post hospitalisation for repeat sampling. Neutrophils were stimulated as previously described with LPS [5 ug/ml] (A) or PMA [100 nM] (B) and NET formation was quantified using SYTOX green. To show linked data from individual patients at acute and follow up time points, fold data were expressed by calculating fold change to DMSO control. Lines link values from the same patient (n=7). There was a significant reduction in LPS induced NET formation at the follow up time point but no difference in PMA stimulated neutrophils. Follow up data were also compared to healthy control data (n=9), these control samples being also used in figure 1A, in response to LPS (C) or PMA (D). LPS induced NETs were significantly higher in follow-up COVID-19 patients compared to healthy controls but there was no difference in PMA stimulated neutrophils. Statistical analysis was performed by a one-tailed paired Student’s t-test (A,B) and a two-tailed unpaired Student’s t-test (C,D) and significance values are as indicated.

## Discussion

Our findings show neutrophils isolated from patients with acute COVID-19 undergo significantly more LPS-induced NETosis than healthy control cells. Although LPS-induced NET formation significantly reduces in COVID-19 patients over time, levels at follow-up time points remained higher than in healthy control cells. Furthermore, NETosis is significantly inhibited by ruboxistaurin *in vitro*. This finding not only supports the importance of the PKC signalling pathway in neutrophils in COVID-19, but also reveals a potential therapeutic strategy for this disease.

Middleton *et al* demonstrated elevated baseline NET levels in neutrophils isolated from COVID-19 patients, which were not further increased by PMA (6). In contrast, we do not show elevated baseline (unstimulated) NET formation, although neutrophils from COVID-19 patients in our study did robustly respond to PMA and generated NETs to levels comparable to healthy control cells. Interestingly, individuals with some of the greatest PMA-induced NET responses went on to require ITU support. Since this was a very small sub-group (n=3) more work is required to determine whether there is an association here.

Whilst others have also shown increased NETosis in people with COVID-19 in response to PMA, a toxin directly activating PKC, we are the first to show an increase in NETs in response to LPS, a receptor-driven neutrophil activator and typically less potent inducer of NETosis. Increased sensitivity to LPS-induced NETosis has implications in the case of secondary infections, and shows neutrophils are primed to increased NET formation to this, and therefore potentially other, proinflammatory stimuli (21). The mechanism by which neutrophils from people with COVID-19 are more sensitive to undergoing NETosis is unclear. SARS-CoV-2 directly triggers NET formation (8) as does sera from COVID-19 patients (9). Increased NET formation in isolated neutrophils *ex vivo* however, suggests this is not as a result of SARS-CoV-2 exposure and is more likely due to the neutrophils being in an activated and primed state and therefore being inherently more sensitive to NET stimuli. This is supported by other studies which describe neutrophil ‘hyperactivation’ in COVID-19, whereby neutrophils are transcriptionally reprogrammed and which is a predictor of severe disease (22-24). Although significantly less than at the acute stage of infection, NET formation in the small subset of individuals available to retest at follow-up time points remained higher several months post hospitalisation than in healthy control cells. This may suggest a pro-NETotic phenotype continues beyond the period of active infection.

Circulating neutrophils from critically ill COVID-19 patients have exaggerated ROS production which may contribute to increased NET production (25). LPS is a weak inducer of NETs in healthy neutrophils compared with PMA, which in part explains why we do not see differences in PMA-NETs when comparing healthy control cells with neutrophils isolated from people with COVID-19 (regardless of the time point). It is possible that upregulation of the TLR4 receptor and/or downstream signalling components in COVID-19 is responsible for the increased sensitivity to LPS-induced (but not PMA-induced) NETs, as seen in monocytes (26). SARS-Cov-2 spike protein activates TLR4 in neutrophil-like cells *in vitro*, and therefore has the potential to cause priming to subsequent exposure to LPS in circulating neutrophils, although whether sufficiently high levels of spike exist in the blood to allow this to occur is unknown (27).

A limitation of our study is around the demographics of the healthy control subjects compared to the patient cohort, the latter of whom are older, have more comorbidities and are receiving medications that may impact on neutrophil function (including dexamethasone). However, studying patients at approximately three months post-hospitalisation means participants serve as appropriate age- and comorbidity-matched controls and allow up to understand differences in neutrophil function at the acute stage of the disease.

Vaccination weakens the link between infection and critical illness, but vaccine breakthroughs are seen, particularly in the case of viral variants which will continue to emerge (28). It is therefore critical that we develop alternative and complementary strategies to prevent severe COVID-19, and the innate immune response is an ideal target for this. Since NETs are known drivers of pathology in a number of diseases including but not limited to COVID-19, targeting NETosis is a logical therapeutic strategy for the future. A growing number of studies describe increased levels of NETosis in COVID-19 as well as the deleterious role of NETs in driving inflammation, thrombosis and disease severity, but few have offered a solution. Our study indicates that ruboxistaurin could reduce NET formation and ultimately diminish airway inflammation and other events including microvascular thrombosis, and is a novel and promising therapeutic strategy for COVID-19 (6, 8). Furthermore, our preliminary data demonstrates reducing NET formation with ruboxistaurin protects airway epithelial cells *in vitro*. Maintaining airway epithelial integrity could provide protection against secondary bacterial infection, which is important since secondary infection is a predictor of death in COVID-19 patients (29, 30). Targeting NETosis in COVID-19 is a strategy shared by others in the field. Therapies inhibiting NET-associated protease activity (NCT04817332) and targeting the breakdown of NETs with DNases are also currently in clinical trials for COVID-19 (NCT04359654). However, ruboxistaurin has the advantage that it prevents NET formation by circulating neutrophils, rather than either modifying or disrupting NETs once they have been formed. Ruboxistaurin has been demonstrated to reduce NET formation in an *in vivo* mouse model and an *in vitro* study of healthy neutrophils, suggesting it has promise in targeting NETs in disease (13, 31). Since phase 3 trials for diabetic retinopathy show ruboxistaurin is a well-tolerated inhibitor of PKC, we believe it could be relatively quick to translate to the clinic, providing a novel therapeutic pathway to treat neutrophil mediated immunopathology in COVID-19 (17).

## Methods

### Human samples

Hospitalised patients with COVID-19 (n=39) admitted to the Royal Hallamshire Hospital, Sheffield, UK were recruited to the study and provided fully informed consent via The Sheffield Teaching Hospitals Observational Study of Patients with Pulmonary Hypertension, Cardiovascular and other Respiratory Diseases (STH-ObS), REC 18/YH/0441, IRAS 248890, project title: Establishing the magnitude, breadth and durability of SARS-CoV-2 induced activation of innate immune blood cells (COVID-19 INNATE)). Ethical approval was given by the Yorkshire & The Humber - Sheffield Research Ethics Committee. Specific project approval was given by the STH-Obs Scientific Advisory Board. All patients had SARS-CoV-2 infection as confirmed by a PCR test. Peripheral blood samples were obtained on average 3 (±3.1 stdev) days post hospital admission. Peripheral blood samples were also obtained for selected patients (n=7) 3-4 months post-hospitalisation at follow-up clinics. Anonymised clinical information was collected for all patients (see Table 1). Blood from healthy volunteers was taken according to the protocol: The control of innate immunity, host-pathogen interactions and leukocyte function in healthy volunteers (REC 05/Q2305/4, STH13927). Ethical approval was given by the Yorkshire & The Humber - Sheffield Research Ethics Committee. All participant information was anonymised and subjects were identified by a unique number.

### Neutrophil isolation

Anti-coagulated blood (10 ml) (1.8 mg/ml EDTA) was processed immediately after phlebotomy and functional studies carried out on freshly isolated cells. Neutrophils were isolated using EasySep™ Direct Human Neutrophil Isolation Kit (Stemcell technologies) as per manufacturer instructions. Mean neutrophil yield was 4.09 × 10^6 ±1.9 cells/ml whole blood.

### SYTOX Green NET assays

Neutrophils were resuspended in RPMI 1640 (without phenol red) and 10 mM HEPES (Thermofisher), and seeded (5 × 10^4) in quadruplicate in a Nunc™ MicroWell™ 96-Well flat bottom plate (Thermofisher). Cells were preincubated with 200 nM ruboxistaurin (Selleckchem) for 1 hour (37°C, 5% CO_2_), before stimulation with either LPS (5 µg/ml) (*Escherichia coli* O111:B4) (Sigma-Aldrich), PMA (100 nM) (Sigma-Aldrich) or DMSO control. Neutrophils were incubated for a further 3 hours before adding SYTOX™ Green nucleic acid stain (555 nM) (Thermofisher) to all wells, to measure extracellular DNA as a surrogate of NET formation. Extracellular DNA was quantified using a fluorescent plate reader (excitation/emission 490/537 nM) and median fluorescence values were reported.

### Immunocytochemistry

Neutrophils (5 × 10^5) were seeded into IBIDI™ µ-slide 8 well chamber slides, pre-incubated for 1 hour with ruboxistaurin (200 nM) and NETosis was induced by LPS and PMA as described above. Cells were fixed with 4% paraformaldehyde (PFA) for 15 minutes at room temperature. Wells were blocked and permeabilized with buffer containing 5% bovine serum albumin (BSA), 0.1% saponin and 5% normal goat serum, and then stained with a rabbit anti-myeloperoxidase (MPO) (A0398) primary antibody for 90 minutes at 37°C. A secondary goat anti-rabbit AlexaFluor® 594 antibody (ab150088) was added for 45 minutes at 37°C. ProLong™ Gold Antifade Mountant with 4’,6-diamidino-2-phenylindole (DAPI) (Thermofisher) was used to stain DNA. Samples were imaged using a NIKON Widefield fluorescence microscope, using the 40x oil immersion objective lens. The DAPI (excitation/emission 395/455 nM) and Texas Red (excitation/emission 555/605 nM) filter sets were used for fluorescent imaging. Images were constructed using FIJI image analysis software and the background was subtracted for the DAPI channel using FiJI.

### Human bronchial epithelial cell culture

Human bronchial epithelial cells (HBEC3-KT) were grown in a humidified incubator at 37°C, 5% CO2. Cells were maintained in basal growth medium; Keratinocyte-SFM (1X) with L-glutamine (Gibco, UK), supplemented with bovine pituitary extract, epidermal growth factor, and Gentamicin sulfate-Amphotericin – 1000 (GA-1000) (Lonza, Switzerland). Cells were passaged twice weekly when at 70-80% confluency and used for experiments between passage 12 and passage 22.

### Cell viability assay

Neutrophils (2.5 × 10^6) were seeded in microcentrifuge tubes and stimulated to induce NET formation with PMA ± ruboxistaurin as described above. Cells were spun at 2500 *g* for 5 minutes and the cell-free supernatant (SPN) were removed and stored at -8°C until required. HBEC3-KT cells were seeded at 1.2 × 10^6 and grown to 90-100% confluency, before overnight incubation in basal media with depleted growth factors. HBEC3-KT cells were incubated with neutrophil SPNs at 1:2 dilution for 24 hours. CellTiter-Glo® was used as a measure of cell viability. Spent media was removed and pre-prepared CellTiter-Glo® reagent added at a 1:2 dilution with basal medium to the tissue culture plate. The plate was incubated (with shaking) at room temperature for 2 minutes then for an additional 10 minutes at room temperature (without shaking). Samples were added in duplicate to a white opaque 96–well plate (Costar) and luminescence determined using a fluorescent plate reader at 480 nM.

## Statistics

Data were plotted and analysed using GraphPad Prism version 9.2. A Shapiro-Wilk normality test was conducted on these data, where n= ⩾6 to determine the use of parametric and non-parametric analyses. Due to missing values a mixed-effect analysis with a Šidák post test was used for comparing NET formation in acute COVID-19 patients with healthy controls. A one-tailed paired t-test was used for comparing NETosis in matched COVID-19 patients at the acute and follow up stage. A one-tailed Wilcoxon matched-pairs signed rank test was used for the DPI inhibition data and a two-tailed Wilcoxon matched-pairs signed rank test was used for the ruboxistaurin inhibition data. A Student’s t-test was used for the epithelial cell viability data and for comparing NET formation in healthy controls and follow up patients.

## Data Availability

The datasets generated during and/or analysed during the current study are available from the corresponding author on reasonable request.

## Author contributions

Experimental work was designed by RD, JC, JM, KLB, RCH, AI, AART, EKT, LRP, IS. Experiments were conducted and data analysis performed by RD, JM, KLB, RCH. Clinical data was provided by JC, AART, CH & JW. Participant recruitment and phlebotomy was performed by JC and AART. Ethics and governance permissions provided by AL. The manuscript was written by RD & LRP. All authors contributed intellectual input to the concept of the study and to the editing and revision of the manuscript drafts.

## Acknowledgments

We thank all of the participants who donated blood to our study. Thanks to Claire Lewis and members of the Sheffield UK-CiC group for helpful discussions. We also thank the Clinical Research Facility staff for support with consent and phlebotomy and Alex Fairman for assistance with follow-up clinics. Our thanks to Darren Robinson at the University of Sheffield Wolfson Light Microscopy Facility for assistance with microscopy and James Chalmers (University of Dundee) for methodological input into the SYTOX green NET assay. This work was supported by University of Sheffield PhD Studentships to RD and JM, The Rosetrees Trust (X/154753-12) (KLB), the Medical Research Foundation (MRF-145-0004-TPG-AVISO) (RCH), the UK Coronavirus Immunology Consortium (UK-CIC) and Medical Research Council AMR cross-council funding to the SHIELD consortium “Optimising Innate Host Defence to Combat Antimicrobial Resistance” (MRNO2995X/1). AART is supported by a BHF Intermediate Clinical Fellowship (FS/18/13/33281).

